# IMpact of a Point-of care UltraSound Examination on the management of acute respiratory or circulatory failure patients in the emergency department: The IMPULSE before-and-after implementation study

**DOI:** 10.1101/2023.09.28.23295699

**Authors:** Sandra Bieler, Damien Tagan, Olivier Grosgurin, Thierry Fumeaux

## Abstract

**Background:** Despite its diagnostic accuracy, the clinical impact of the use of Point-of-care Ultrasound (POCUS) in the emergency department (ED) is not well described, especially when performed by junior in-training residents.

**Aim of the study:** to assess the effect of a short, structured POCUS training program on the management of ED patients by in-training residents.

**Method:** IMPULSE is a before-and-after implementation study, evaluating the impact of a structured POCUS training program for ED in-training residents on the management of patients admitted for acute respiratory and/or circulatory failure in a Swiss regional hospital. The training curriculum was organized in three stages and combined an on-line training course, an 8-hour practical hands-on session, and 10 supervised POCUS exams. The ED residents who successfully completed the curriculum participated in the study. Observed outcomes were time to ED diagnosis, rate of correct diagnosis made during the ED stay and time needed to reach it, time to prescribe an appropriate treatment, and hospital mortality. Standard statistical analyses were performed with the use of Chi-square and Mann-Whitney U tests as appropriate, completed by a Bayesian analysis, with a Bayes Factor (BF) > 3 considered as significant.

**Results:** Sixty-nine patients were included before the training program implementation and 54 after. After implementation, the median time to ED diagnosis was 25 minutes (IQR 44) vs 30 minutes (IQR 56) before implementation, a difference that was significant (BF 9.6). The rate of correct diagnosis was higher (95 vs 52 %) (p<0.001) and the time to make this correct diagnosis was significantly shorter after implementation (25 minutes, IQR 45, vs 43, IQR 60) (BF 5.0). This had an impact on the median time to prescribe the appropriate therapy, with a trend toward a shorter delay (47 minutes, IQR 76, vs 70, IQR 100) (BF 2.0). Eventually, there was a significant difference in hospital mortality (13 % vs 5.5 %, BF 15.7).

**Conclusion:** the IMPULSE study shows that the implementation of a short, structured POCUS training program for ED residents to use ultrasound for the initial evaluation of acute respiratory and circulatory failure patients has an impact on diagnostic accuracy, on time to make a correct diagnosis and to prescribe an appropriate therapy, and eventually possibly on hospital mortality. If these results are reproduced in other settings, POCUS use by ED residents after short, structured training curriculum could become the standard of care for these patients.

## BACKGROUND

Acute respiratory failure (ARF) and acute circulatory failure (ACF) are frequent causes of ED admissions and are associated with a significant morbidity and mortality, and a high level of ED resource consumption. Timely and adequate management can reduce these consequences but depends on an efficient diagnosis workup^1^. Traditionally, this work-up is guided by history taking and physical examination, which have been shown to be inaccurate in the ED^2-4^. Basic laboratory and imaging procedures are often completed with more advanced modalities, such as transthoracic echocardiography or computed tomography, at the cost of increased duration of ED stay, resource consumption, and potential adverse events^5-7^. Point of care ultrasound (POCUS) performed by non-radiologists or non-cardiologists is a bedside non-invasive diagnostic tool that has been shown to be very accurate to identify the etiologic cause of ARF or ACF, with no significant side effects ^8-20^. POCUS is now included in many educational programs of emergency physicians^21-27^. Nevertheless, there is still no strong evidence that the diagnostic accuracy of POCUS translates into a clinically relevant difference in patients outcomes^18,28-33^. Moreover, in most of the published studies, POCUS was performed by trained experts, who were not directly in charge of the patient and often blinded to clinical data. The diagnostic accuracy of POCUS when performed by less experienced first-contact ED physicians after a short, structured training period is not therefore well described. Despite these limitations, the recent clinical guideline from the ACP (American College of Physicians) recommends the use of POCUS in addition to standard diagnostic processes in patients with acute dyspnea, while awaiting the results of ongoing randomized studies^34-35^.

We designed the <u>**IM**</u>pact of a <u>**P**</u>oint-of care <u>**Ul**</u>tra<u>**S**</u>ound <u>**E**</u>xamination (IMPULSE) study to clarify some of these unresolved issues, by studying the impact of a structured POCUS training program for ED in-training residents on several outcomes of ED patients admitted for ACF and/or ARF. The design of a before-and-after implementation study was selected to avoid the methodological problems associated with blinding and randomization in a single-center study^35^.

## STUDY DESIGN AND METHODS

### Study design and intervention description

IMPULSE is a single center before-and-after observational implementation study of a structured POCUS training program for ED residents of a regional hospital (*Hôpital de Nyon*, Switzerland). During the pre-implementation period (phase 1), the management of patients was not altered, and POCUS could be performed by trained senior physicians, as part of the standard management work-up since 2010. During the intervention phase, all the residents who were planned to work in the ED during the post-implementation period (phase 2) were included in the program developed by the AURUS (*Association des Urgentistes et Réanimateurs intéressés à l’Ultrasonographie*), with a content that is in line with the ESICM consensus document^36-38^. This training curriculum was organized in three stages:

- a 20-hour online course (“www.pocus.academy“)on general principles of ultrasonography, theoretical and practical aspects of image acquisition and interpretation, in transthoracic cardiac, vascular, lung, and abdominal POCUS^39^. The module includes a formal evaluation of knowledge through a multiple-choice questionnaire, which must be completed to progress to the next step
- an 8-hour practical hands-on session, with POCUS examinations performed on healthy volunteers and on simulators, in groups of 3 students supervised by one trainer, focused on the technical aspects of obtaining interpretable images. The session includes a formal assessment of the image acquisition and interpretation skills. This assessment is mandatory to proceed to the next step.
- the practice of at least 10 directly supervised POCUS full examinations performed in real conditions in the ED, with a formal evaluation of the ability to acquire and interpret good quality images and to integrate them into the clinical management.

At the end of the training process, the residents fulfilling all training objectives were allowed to participate in the phase 2. A *Sparq Ultrasound System® (*Philips AG Healthcare, Horgen, Switzerland) was used for all POCUS exams, which were realized with a linear 4-12 MHz probe and a phased array 1-4 MHz probe. The POCUS was encouraged to be performed as soon as possible for all included patients, following a standardized protocol assessing 19 specific sonographic signs (e-Figure 1, supplemental material), looking for signs suggesting pulmonary embolism, left heart failure, hypovolemic state, tamponade, pneumonia, pneumothorax, or an abdominal condition. All POCUS images were recorded, and a standardized report form was to be filled by the resident (e-Figure 2, supplemental material). All images were mandatorily supervised - directly or afterwards - by a senior physician trained in POCUS, to confirm the findings reported in the standardized form.

All other diagnostic procedures were used at the discretion of the clinician, including basic POCUS performed by the attending physician, and advanced US examination by a fully trained radiologist or cardiologist.

### Inclusion and exclusion criteria

In both phases, all consecutive adult patients (≥ 18 years) presenting with ARF and/or ACF were screened to be included in the study. ARF was defined by the presence of either signs of respiratory distress or a respiratory rate over 20/min, and oxygen saturation at pulse oximetry < 92 % on room air or the necessity to administer oxygen to maintain a saturation of ≥ 92 %. ACF was defined by the presence a systolic blood pressure < 90 mmHg, and clinical signs of hypoperfusion signs (agitation or consciousness alteration, skin mottling, oligo-anuria) or hyperlactatemia (> 2.0 mmol/l).

Exclusion criteria were a known or an immediate diagnosis (such as STEMI or referral for an externally identified diagnosis), the need for an immediate life-saving procedure (such as cardiopulmonary resuscitation), trauma, palliative care, and patients care refusal.

To preserve the organization of the ED and to favor the inclusion of patients for whom uninterrupted management seemed likely, final inclusion of patients and start of observation was left to the discretion of the resident in charge, based on his evaluation of the ED situation and workload.

The study was approved by the *Commission Cantonale d’Ethique* from the *Canton de Vaud* (CER-VD, protocol number 194/15). Due to the observational design of the study, and as the practice of POCUS was already part of the usual care in the ED of the institution, a signed individual informed consent was required only for the use of the data collected for the study. Consequently, in order not to delay the management of the patients, a short oral information was given to the patient at the start of the observation. A full information about the study was then given to the patient as soon as possible. Inclusion was finalized after individual signed consent. In the case of patient refusal to participate, all study material was destroyed.

### Data collection

During both phases, only one ED resident at a time was participating in the study. The ED resident was asked to report in a specific form the exact time of the start of the observation, the time at which the diagnosis was made and the time at which the specific treatment was prescribed. In the report form, a list of diagnosis and therapies was proposed (e-Figure 1, supplemental material). The ED resident was equipped with a back-up audio recorder, which was started at the initial contact with the patient. All recordings were maintained confidential to the investigators only, who analyzed them to check the written data. Based on these data, time to make a diagnosis, to prescribe a targeted appropriate treatment, and ED length of stay were computed, and rounded to 5 minutes intervals. The hospital discharge summary was retrospectively analyzed to compare the diagnosis made during the ED stay with the final hospital diagnosis and to assess the hospital mortality.

### Statistical analysis

All data were analyzed with the free open-source JASP tool^40^. Median values and IQR are reported for descriptive statistics of continuous variables, and absolute numbers and proportion for categorical variables. Differences in proportions of categorical variables between phases were analyzed by Chi-square test, with a significant level set at p values < 0.05. Differences in continuous variables and time intervals between phases were analyzed with a Mann-Whitney U test, completed by a Bayesian approach. For this analysis, the alternative hypothesis was that the time intervals would be greater in phase 1 than in phase 2, with a prior described by a Cauchy distribution centered around zero and with a width parameter of 1.00. This width parameter was chosen after an equivalence Bayesian independent samples T-test analysis and corresponds to a probability of 50% that the effect size lies between -1.000 and 1.000. The statistical significance of Bayesian analysis was expressed with Bayes factor (BF), a value between 3 and 10 being considered moderate evidence, and a value over 10 representing strong evidence. For hospital mortality comparison between the two phases, a Bayesian analysis was also performed, with an independent binomial analysis, with fixed rows.

## RESULTS

### Patients

Among 139 eligible patients, 123 with ARF (117) and/or ACF (20) were included in the final analysis: 3 patients refused to participate afterwards, 1 patient did not have the inclusion criteria, and data were missing for 12 patients (Figure 1). Phase 1 lasted from September 4, 2015, to May 28, 2016 (268 days) and included 69 patients (1 patient every 4 days). Implementation of the POCUS training phase lasted from May 29, 2016, to September 14, 2016. During this period, twelve residents were successfully trained. Phase 2 lasted from September 15, 2016, to February 7, 2018 (511 days), and included 54 patients (1 patient every 9 day). The median age of the included patients was 77, and most of the patients were included for respiratory distress and hypoxemia. The admission characteristics of the included subjects are representative of this category of ED patients (Table 1).

**Figure 1.**
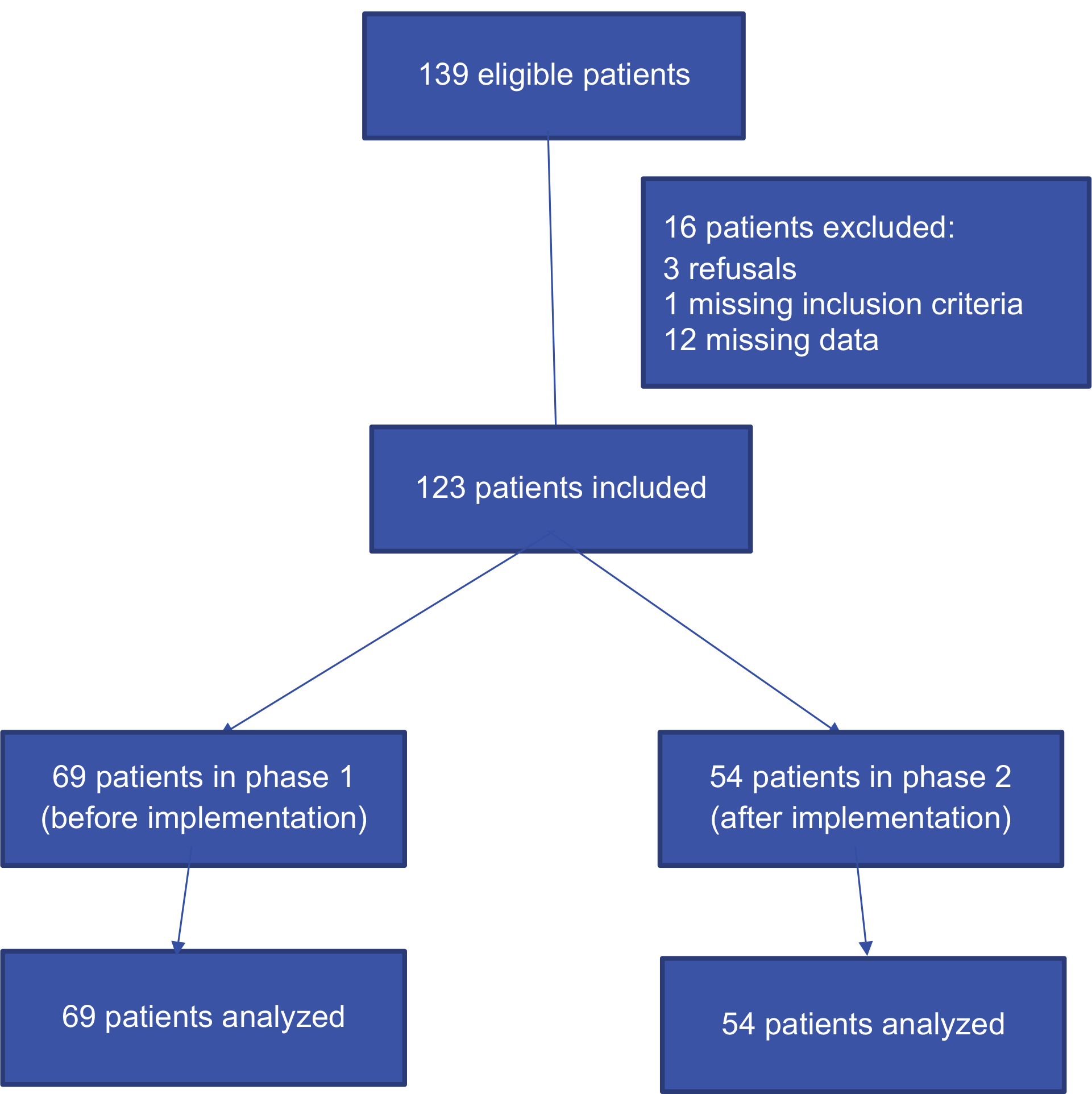
Study flow chart CONSORT.

**Table 1.**
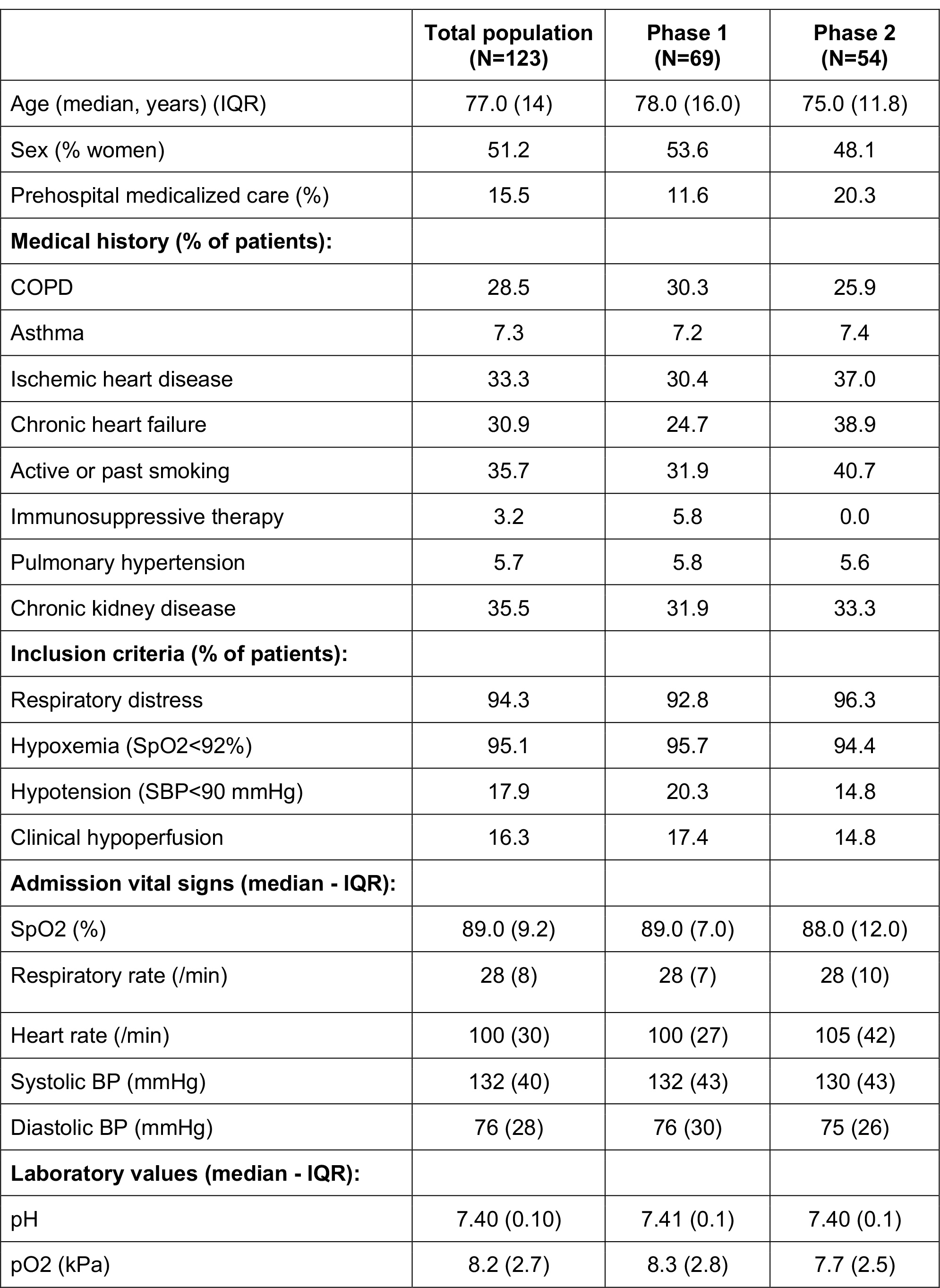
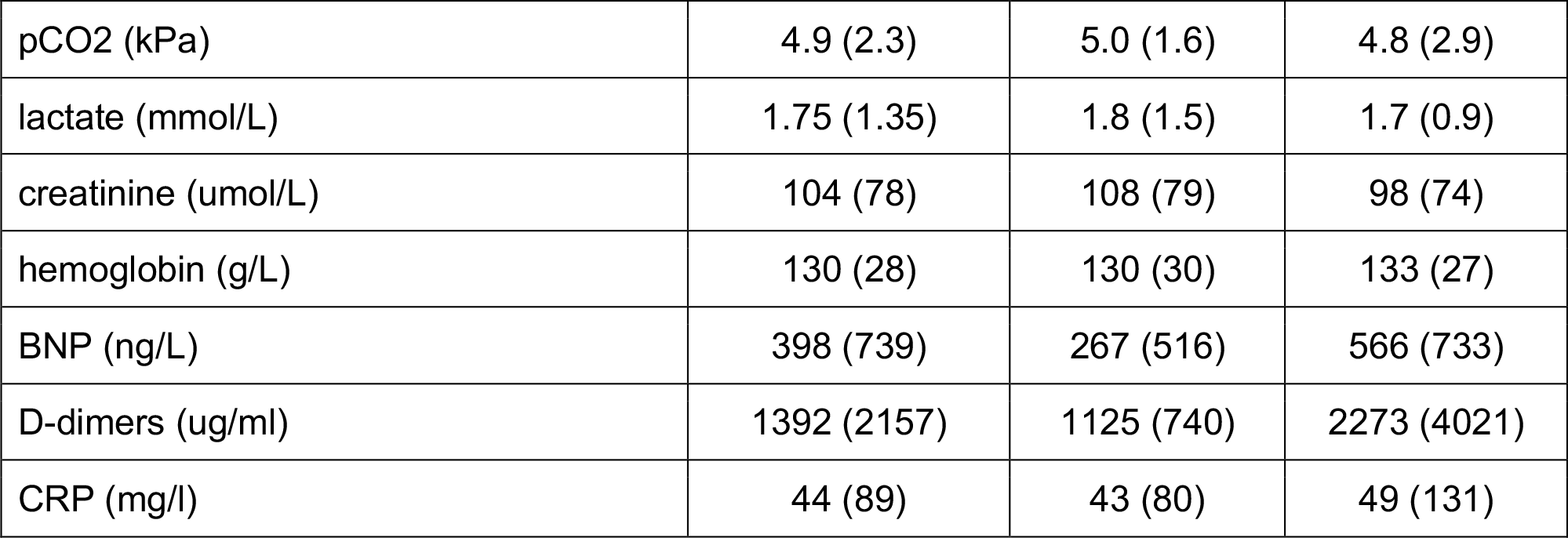
Patients characteristics at admission.

|  | **Total population**<br>**(N=123)** | **Phase 1**<br>**(N=69)** | **Phase 2**<br>**(N=54)** |
| --- | --- | --- | --- |
| Age (median, years) (IQR) | 77.0 (14) | 78.0 (16.0) | 75.0 (11.8) |
| Sex (% women) | 51.2 | 53.6 | 48.1 |
| Prehospital medicalized care (%) | 15.5 | 11.6 | 20.3 |
| **Medical history (% of patients):** |  |  |  |
| COPD | 28.5 | 30.3 | 25.9 |
| Asthma | 7.3 | 7.2 | 7.4 |
| Ischemic heart disease | 33.3 | 30.4 | 37.0 |
| Chronic heart failure | 30.9 | 24.7 | 38.9 |
| Active or past smoking | 35.7 | 31.9 | 40.7 |
| Immunosuppressive therapy | 3.2 | 5.8 | 0.0 |
| Pulmonary hypertension | 5.7 | 5.8 | 5.6 |
| Chronic kidney disease | 35.5 | 31.9 | 33.3 |
| **Inclusion criteria (% of patients):** |  |  |  |
| Respiratory distress | 94.3 | 92.8 | 96.3 |
| Hypoxemia (SpO<sub>2</sub><92%) | 95.1 | 95.7 | 94.4 |
| Hypotension (SBP<90 mmHg) | 17.9 | 20.3 | 14.8 |
| Clinical hypoperfusion | 16.3 | 17.4 | 14.8 |
| **Admission vital signs (median - IQR):** |  |  |  |
| SpO<sub>2</sub> (%) | 89.0 (9.2) | 89.0 (7.0) | 88.0 (12.0) |
| Respiratory rate (/min) | 28 (8) | 28 (7) | 28 (10) |
| Heart rate (/min) | 100 (30) | 100 (27) | 105 (42) |
| Systolic BP (mmHg) | 132 (40) | 132 (43) | 130 (43) |
| Diastolic BP (mmHg) | 76 (28) | 76 (30) | 75 (26) |
| **Laboratory values (median - IQR):** |  |  |  |
| pH | 7.40 (0.10) | 7.41 (0.1) | 7.40 (0.1) |
| pO<sub>2</sub> (kPa) | 8.2 (2.7) | 8.3 (2.8) | 7.7 (2.5) |
| pCO<sub>2</sub> (kPa) | 4.9 (2.3) | 5.0 (1.6) | 4.8 (2.9) |
| lactate (mmol/L) | 1.75 (1.35) | 1.8 (1.5) | 1.7 (0.9) |
| creatinine (umol/L) | 104 (78) | 108 (79) | 98 (74) |
| hemoglobin (g/L) | 130 (28) | 130 (30) | 133 (27) |
| BNP (ng/L) | 398 (739) | 267 (516) | 566 (733) |
| D-dimers (ug/ml) | 1392 (2157) | 1125 (740) | 2273 (4021) |
| CRP (mg/l) | 44 (89) | 43 (80) | 49 (131) |

### General ED management (Table 2)

The median ED stay duration was 235 minutes (IQR 120). During ED stay, 80 % of the patients had a chest X-ray, one third a chest CT-scan, and 38 % had a POCUS performed by a senior supervisor. Pneumonia was the most frequent diagnosis, followed by acute heart failure. Antibiotics and diuretics were the most frequently prescribed therapies during ED stay. Except for two patients (one death and one home discharge), all patients were hospitalized, in around half of the cases in the Intensive Care Unit (ICU) (Table 2).

**Table 2.** ED management.

|  | **Total population**<br>**(N=123)** | **Phase 1**<br>**(N=69)** | **Phase 2**<br>**(N=54)** |
| --- | --- | --- | --- |
| **Imaging (%):** |  |  |  |
| Chest X-ray | 79.7 | 94.2 | 61.1 |
| Thoracic CT | 33.3 | 30.4 | 37.0 |
| Abdominal CT | 11.4 | 7.2 | 16.7 |
| Abdominal US | 3.3 | 5.8 | 0.0 |
| Transthoracic echocardiography | 2.4 | 2.9 | 1.9 |
| POCUS by senior physician | 38.2 | 34.8 | 42.6 |
| **ED Diagnosis (%):** |  |  |  |
| Pneumonia | 34.1 | 37.7 | 29.6 |
| Acute heart failure | 33.3 | 27.5 | 40.7 |
| Acute exacerbation of COPD | 10.6 | 13.0 | 7.4 |
| Non pulmonary sepsis | 8.9 | 11.6 | 5.6 |
| Pulmonary embolism | 4.1 | 1.4 | 7.4 |
| Pericardial effusion | 2.4 | 0.0 | 5.6 |
| Cardiogenic shock | 1.6 | 1.4 | 1.9 |
| Other diagnosis | 5.0 | 7.4 | 1.8 |
| **Specific ED therapies (%)\*:** |  |  |  |
| Antibiotics | 52.0 | 56.5 | 37.0 |
| Diuretic therapy | 39.8 | 34.8 | 46.3 |
| Bronchodilators | 21.9 | 26.1 | 16.7 |
| Noninvasive ventilation | 20.3 | 21.7 | 18.5 |
| Steroids | 13.8 | 14.5 | 13.0 |
| Anticoagulation | 11.4 | 7.2 | 16.7 |
| Vasopressors | 9.8 | 8.7 | 11.1 |
| **Patient destination after ED stay (%):** |  |  |  |
| Ward | 47.2 | 52.2 | 40.7 |
| ICU | 47.2 | 43.5 | 51.9 |
| Other hospital | 4.0 | 2.9 | 5.6 |
| Home | 0.8 | 1.5 | 0.0 |
| Death in the ED | 0.8 | 0.0 | 1.8 |

\* Some patients may have received more than one therapy

### Comparison between phase 1 and phase 2

Compared to phase 1, there was a significant decrease in the median time to final ED diagnosis in phase 2 (30 (IQR 56) vs 25 minutes (IQR 44) - BF 9.6) (Table 4), a 5-minute difference that may be clinically relevant in emergency situations. The rate of confirmed ED diagnosis during the hospital stay was 52 % during phase 1 and 94 % in phase 2, a difference that is highly significant (Chi-squared test 26.146, *P* < 0.001) (Table 3). These results show that the high diagnostic accuracy of POCUS is preserved when performed in real-life conditions by ED residents, after a short, structured training. When the ED diagnosis was later confirmed during the hospital stay, the time needed to make this diagnosis was significantly reduced in phase 2 (25 (IQR 45) vs 43 minutes (IQR 60) - BF 5.0), a 18-minute difference that is only moderately significant in the Bayesian analysis but clinically highly relevant. Finally, the time to order the appropriate therapy was reduced from 70 minutes (IQR 100) in phase 1 to 47 minutes (IQR 76) in phase 2 (BF 2.0). Although limited by the power of the study, these results suggest that POCUS use may not only be associated with a higher rate of correct diagnosis in the ED, but also with a quicker diagnostic and therapeutic process. A decrease in the duration of ED stay was also observed, significant in Bayesian analysis, although probably not clinically relevant (Table 4). Eventually, the hospital mortality was reduced during phase 2 (6 % vs 13 % in phase 1), a difference that was very significant in a Bayesian analysis (BF 16.04) (Table 5).

**Table 3.** Confirmation of ED diagnosis during hospital diagnosis. - Contingency table

| **Phase** |  | **Diagnostic confirmed during hospital stay** |  | **Total** |
| --- | --- | --- | --- | --- |
|  |  | **No** | **Yes** |  |
| 1 | Count | 33 | 36 | 69 |
|  |  | 48 % | 52 % |  |
| 2 | Count | 3 | 51 | 54 |
|  |  | 6 % | 94 % |  |
| Total | Count | 36 | 87 | 123 |
|  |  | 29 % | 71 % |  |

Chi-squared 26.146 - $P < 0.001$

**Table 4.** ED time intervals (minutes) (median - IQR)

|  | Phase 1<br>(N= 69) | Phase 2<br>(N=54) | BF | *P* value |
| --- | --- | --- | --- | --- |
| Time to final diagnosis | 30 (56) | 25 (44) | 9.56 | 0.327 |
| Time to final diagnosis confirmed during stay | 43 (60) | 25 (45) | 5.02 | 0.327 |
| Time to administer targeted therapy | 70 (100) | 47 (76) | 1.96 | 0.311 |
| Duration of ED stay | 238 (137) | 230 (115) | 4.18 | 0.415 |

BF: Bayes Factor - Alternative hypothesis phase 1 > phase 2 - prior: Cauchy, scale 1.0 *P* value calculated with Mann-Whitney U test

**Table 5.**
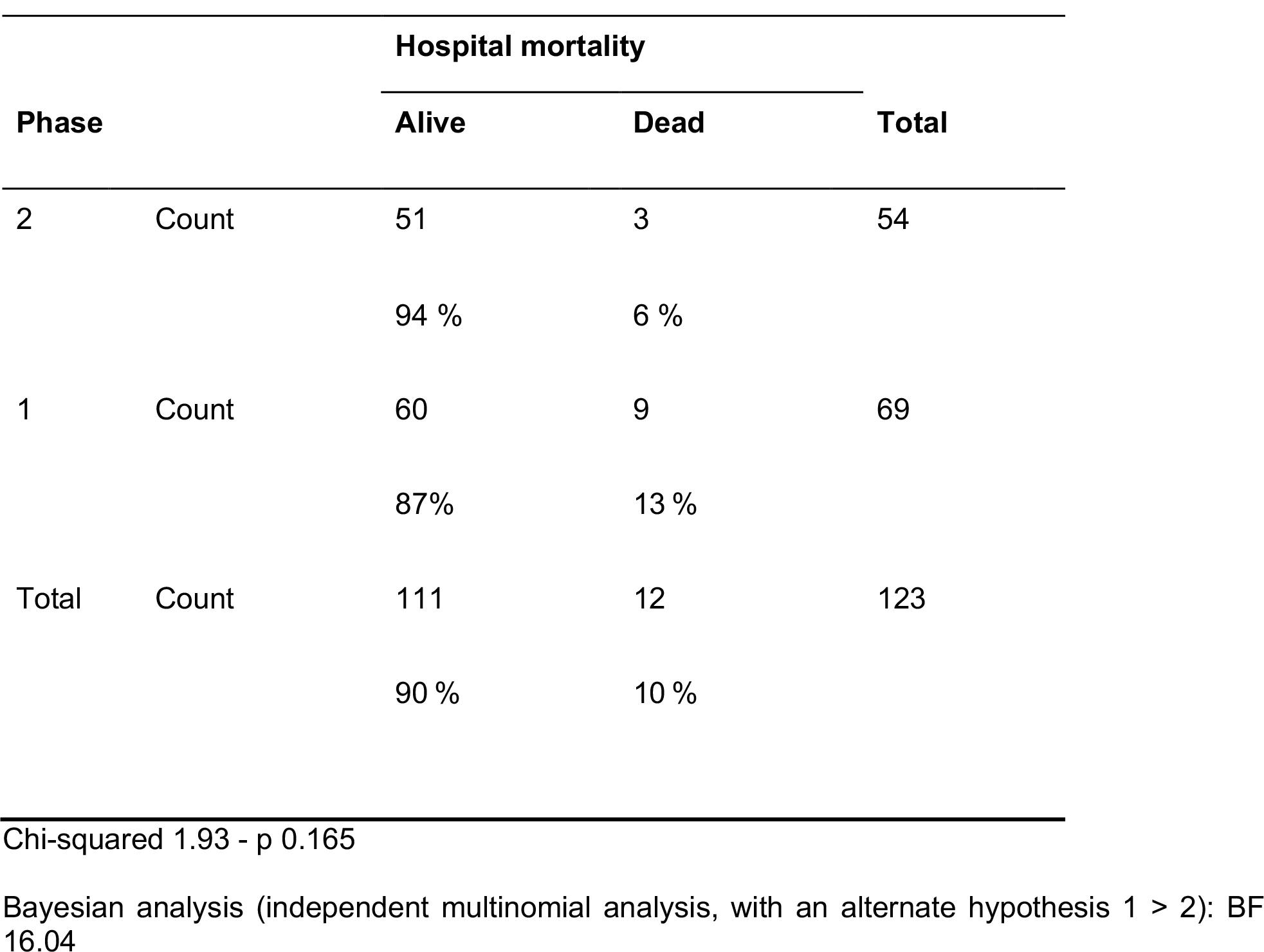
Hospital mortality. Contingency table

|  |  | Hospital mortality |  |  |
| --- | --- | --- | --- | --- |
| Phase |  | Alive | Dead | Total |
| 2 | Count | 51 | 3 | 54 |
|  |  | 94 % | 6 % |  |
| 1 | Count | 60 | 9 | 69 |
|  |  | 87% | 13 % |  |
| Total | Count | 111 | 12 | 123 |
|  |  | 90 % | 10 % |  |

Chi-squared 1.93 - p 0.165
Bayesian analysis (independent multinomial analysis, with an alternate hypothesis 1 > 2): BF 16.04

Due to the small population sample, we did not perform a formal statistical analysis of the characteristics of the patients, the components of ED management, the diagnosis distribution and the administered therapies (Table 1 and 2). Nevertheless, we evidenced a clear decrease in the number of chest X-ray examinations performed during phase 2, with an increase in the number of CT-scan performed during the ED stay. During phase 1, a POCUS was performed by senior supervisors, per study design, in 35 % of the patients, as during phase 2, all patients had a POCUS performed by the resident in charge, a second POCUS being performed by the senior supervisor in almost half of the case (Table 2).

## DISCUSSION

The objective of IMPULSE was to show that the implementation of a short, structured training program for ED residents could have an impact on the management and outcome of patients admitted for ARF or ACF. A before-and-after implementation design was chosen to be as close as possible to a randomized trial, while avoiding the problems of the contamination bias between the two groups. The POCUS training curriculum (AURUS) was selected as it was already well established in the institution, and because it is in line with the updated recommendations concerning the training objectives of the current guidelines^38,39^.

Our results show that implementation of the structured AURUS POCUS program is associated with a significantly higher rate of diagnostic accuracy, with shorter time to make a diagnosis, a difference that is greater when the ED diagnosis is later confirmed during the hospital stay. The decreased mortality is to be taken with caution, as the design of the study, the small sample size with differences in the case mix expose to multiple potential bias. This effect on mortality should therefore be reproduced in other studies. As far as we know, these data are the first to show a clinical impact of the use of POCUS in the ED and to support a structured training protocol for in-training ED physicians. If the IMPULSE results are reproduced, the first line use of POCUS combined with history taking and clinical examination, just like an enhanced stethoscope, could become a standard of care. Another recent publication similarly suggested that the use of POCUS by physicians of various levels of experience was associated with improved administration of appropriate therapies, even though diagnostic accuracy was not improved^41^.

The IMPULSE study has several strengths. The study design reflects real-life conditions of the majority of ED, where patients are initially taken in charge by junior physicians, under the supervision of experienced senior emergency specialists. The characteristics of the included patients and the diagnosis that were made in the ED show that this study sample is representative of the population of interest for the use of POCUS, with a significant associated morbidity and mortality, for whom a significant impact on the outcome can be evidenced even in a small sample. The reported intervention, which is the implementation process of a structured POCUS training and not only the use of POCUS by experts, is described for the first time in a real-life context, showing not only that it is feasible, but also that it has an impact on the management and outcome of patients. It moreover avoids the contamination bias seen in several studies. The use of POCUS in phase 1 reflects the practice of most ED departments, where POCUS is usually only performed by senior physicians, often later in the management of the patients. The high rate of inaccurate diagnosis during this phase reflects the diagnostic challenge in the ED^42-44^. Another strength of the IMPULSE study is the signal of a clinically relevant impact on the patient outcome: ideally, morbidity and mortality should be the endpoint of choice for interventional studies in ED patients, but as POCUS is not a therapeutic procedure, the effect on outcome can only be driven by a quicker and more appropriate administration of efficient therapies. IMPULSE shows that POCUS not only improves diagnostic accuracy, but shortens the time to make a correct diagnosis, particularly when the diagnosis is correctly made, and that this reduction may be associated with a decrease in hospital mortality.

Despite our efforts to use the most appropriate methodology, the IMPULSE study has several limitations that should be considered. First, it is not a randomized trial, but randomization of patients in two parallel groups can lead to contamination bias between arms. A cluster randomization may decrease this risk but not suppress it. We therefore chose the before-and-after implementation design as the best way to get a quasi-randomization of the patients without the risk of contamination. The limited sample of included patients, in a single center, is another important limitation. Despite a long period of recruitment, the number of patients included in the study was low, particularly in phase 2, with one included patient every 9 days, suggesting that not all consecutively admitted patients were included. This is mostly due to the difficulty of conducting single center studies in small structures, without devoted clinical study resources. Despite this important limitation, we nevertheless think that the patients are not only representative of the usual clinical activity of our ED but are also comparable to the usual patients admitted for ARF and ACF in most ED around the world, as demonstrated by their characteristics and the corresponding diagnosis. Of course, our results should be reproduced in other clinical settings, with the inclusion of a larger sample of patients, before any firm conclusion can be made on the impact of POCUS when used as first-line examination by in-training ED residents. These limitations do not alter the fundamental message of the results presented herein.

Some secondary findings of the IMPULSE study also deserve some attention. During phase 2, even though POCUS was performed by first-line residents, a senior physician repeated the examination in almost half of the cases, a proportion that is greater than the one third of patients who had a POCUS during phase 1 (Table 2). This may have been for verification purposes, but another hypothesis may be that the practice of POCUS by junior physicians is a trigger for more experienced physicians to also perform it more frequently, and not only for supervision. Similarly, and although this should be interpreted with caution, we evidenced a reduction in the number of chest X-ray during phase 2 (performed in 61 % of patients only), suggesting that POCUS may be used in place of this poorly performing exam. At the same time, the number of CT-scan exams was higher during phase 2, and this may be interpreted in two different ways: it could be a negative effect of POCUS, driving supervisors to make more CT-scans to confirm or reject a possible diagnosis made by their inexperienced junior colleagues. The fact that the number of POCUS exams performed by supervisors also increased suggests that this may be a more positive effect, POCUS giving a better assessment of the clinical situation and driving a more appropriate use of advanced diagnostic modalities. Similar studies published in the future will probably address these findings and may confirm these trends, while providing clarification as to the causes of these increases in CT-scan use.

## CONCLUSION

The IMPULSE study shows that a short, structured training program for ED residents is feasible and allows them to use POCUS in the first line management of patients with ARF or ACF. The use of POCUS by these less experienced physicians is associated with a diagnostic accuracy comparable to published data, with a decrease in the time needed to make a correct diagnosis, and with a possible increase in the prescription of an appropriate therapy and a decrease in hospital mortality. The results of IMPULSE also validate the AURUS training curriculum, showing that this structured stepwise approach of training is not only feasible, but efficient. These results need to be reproduced and confirmed in other settings with larger patient samples, but the methodology presented herein is appropriate to limit the problems of blinding and randomization in the study of such diagnostic tools and may be used by future studies.

## Data Availability

All data produced in the present study are available upon reasonable request to the authors

## Abbreviations

ACF: Acute circulatory failure
ARF: Acute respiratory failure
BF: Bayes Factor
CER-VD: Commission Cantonale d’Ethique from the Canton de Vaud
ED: Emergency department
ICU: Intensive care unit
IMPULSE: Impact of a point of care examination
IQR: Interquartile range
POCUS: Point-of-care ultrasound

## SUPPLEMENTAL MATERIAL

**e-Figure 1.**
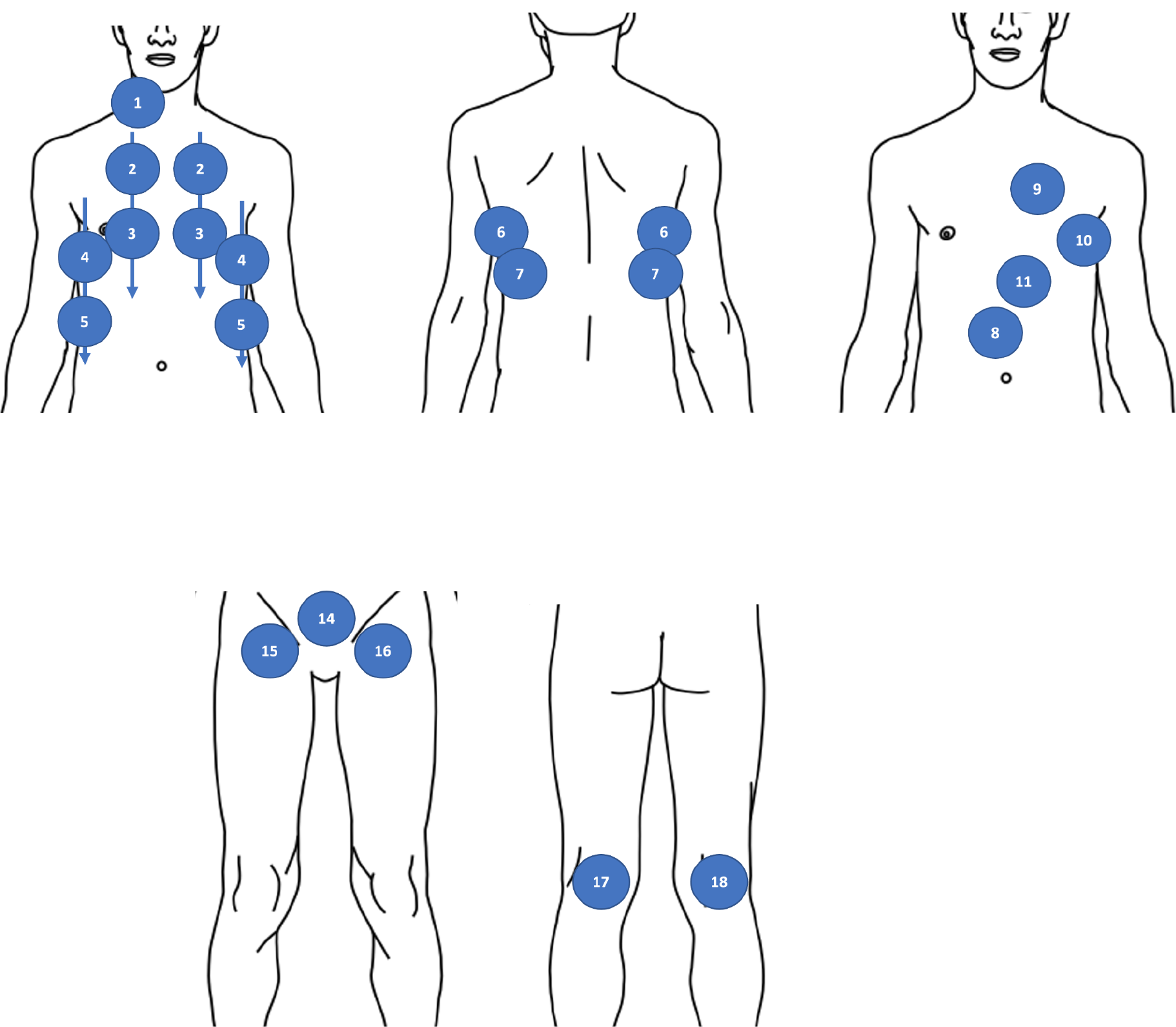
POCUS protocol. 1.internal jugular vein 2-5. anterior pulmonary view/anterior axillary line view 6-7. posterobasal pulmonary view 8. inferior vena cava 9. parasternal short and long axis cardiac view 10. apical four cavities cardiac view 11. subcostal cardiac view 12. hepato-renal space 13. spleno-renal space 14. supra-pubic view 15-18. femoro-popliteal veins

**e-Figure 2.**
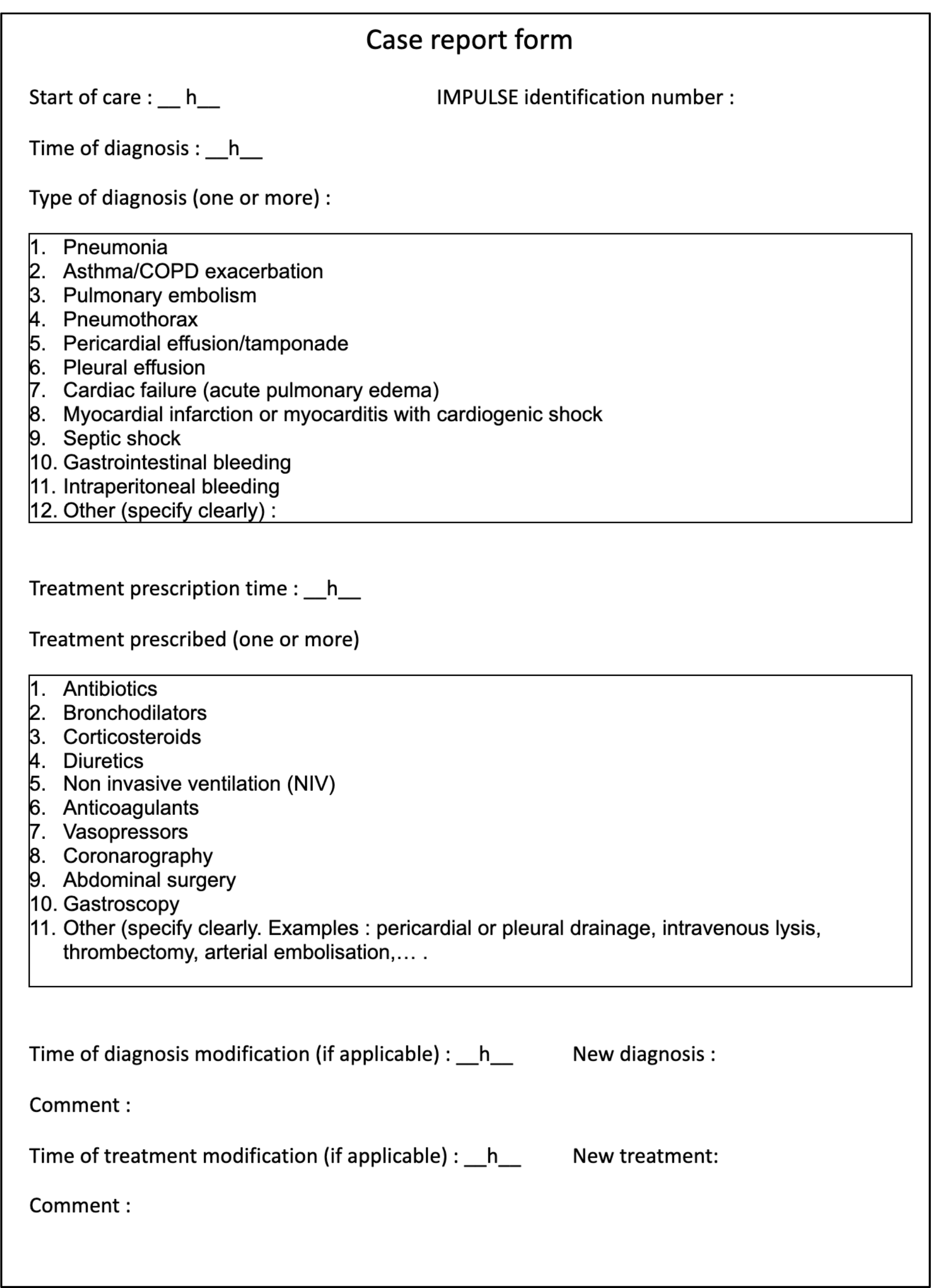
Case report form (adapted from original form in French)

