## Supplemental tables for "IMpact of a Point-of care UltraSound Examination on the management of acute respiratory or circulatory failure patients in the emergency department: The IMPULSE before-and-after implementation study"

#### SUPPLEMENTAL MATERIAL

##### e-Figure 1 - POCUS protocol

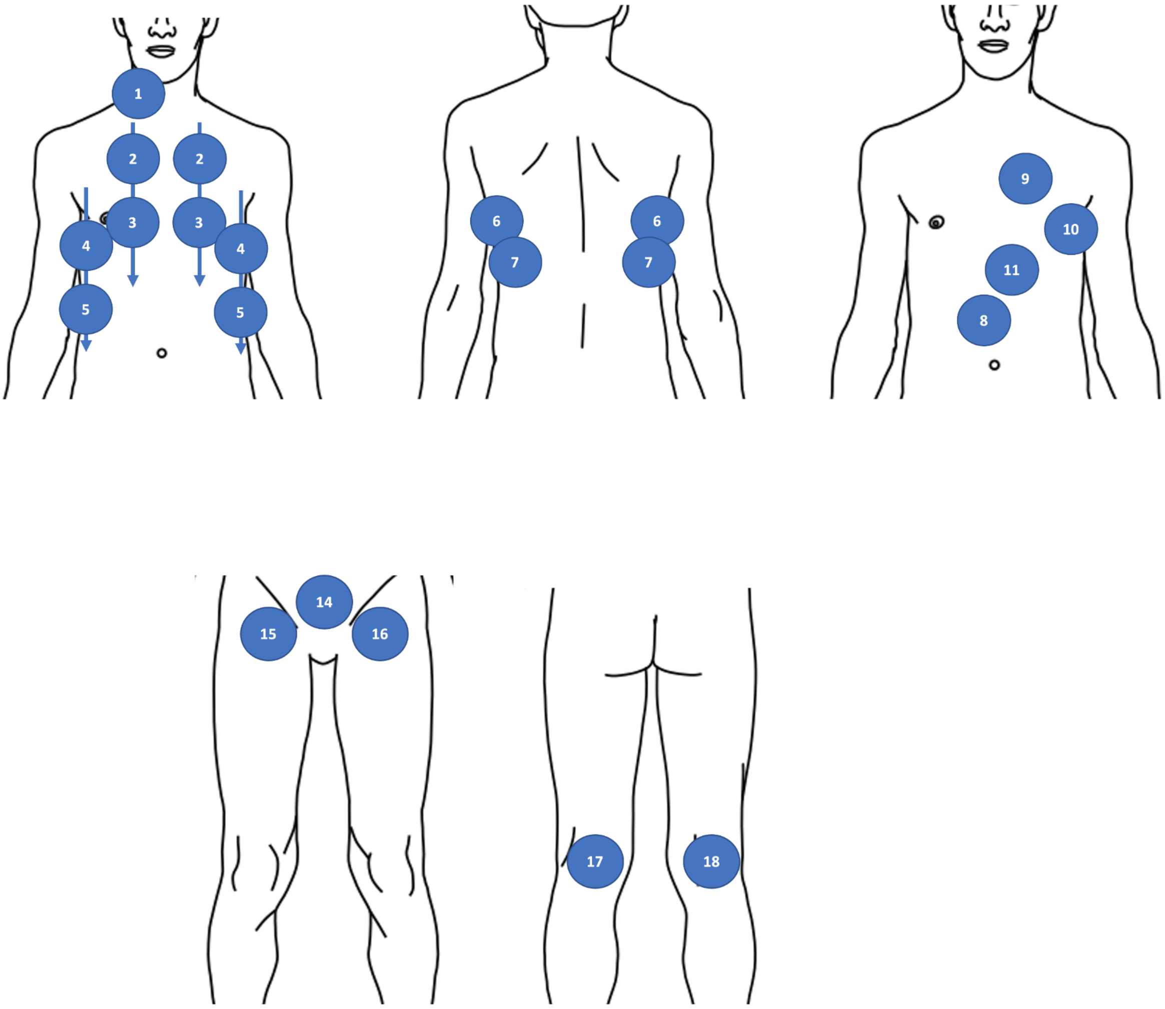


1.internal jugular vein 2-5. anterior pulmonary view/anterior axillary line view 6-7. posterobasal pulmonary view 8. inferior vena cava 9. parasternal short and long axis cardiac view 10. apical four cavities cardiac view 11. subcostal cardiac view 12. hepato-renal space 13. spleno-renal space 14. supra-pubic view 15-18. femoro-popliteal veins

**e-Figure 2 - Case report form (adapted from original form in French)**

### Case report form

Start of care : __ h__ IMPULSE identification number :

Time of diagnosis : __h__

Type of diagnosis (one or more) :

1. Pneumonia
2. Asthma/COPD exacerbation
3. Pulmonary embolism
4. Pneumothorax
5. Pericardial effusion/tamponade
6. Pleural effusion
7. Cardiac failure (acute pulmonary edema)
8. Myocardial infarction or myocarditis with cardiogenic shock
9. Septic shock
10. Gastrointestinal bleeding
11. Intraperitoneal bleeding
12. Other (specify clearly) :

Treatment prescription time : __h__

Treatment prescribed (one or more)

1. Antibiotics
2. Bronchodilators
3. Corticosteroids
4. Diuretics
5. Non invasive ventilation (NIV)
6. Anticoagulants
7. Vasopressors
8. Coronarography
9. Abdominal surgery
10. Gastroscopy
11. Other (specify clearly. Examples : pericardial or pleural drainage, intravenous lysis, thrombectomy, arterial embolisation,… .

Time of diagnosis modification (if applicable) : __h__ New diagnosis :

Comment :

Time of treatment modification (if applicable) : __h__ New treatment:

Comment :
